# Trigeminal Nerve Blocks In Refractory Trigeminal Neuralgia: About 21 Cases Reported At The Limoges University Hospital

**DOI:** 10.1101/2020.10.23.20218362

**Authors:** Nicolas Jacques, Simon Karoutsos, Loïc Marais, Nathalie Nathan-Denizot

## Abstract

**Introduction:** Despite limited scientific evidence, trigeminal nerve blocks are alternative therapies for refractory trigeminal neuralgia (RTN). The duration of analgesia far exceeds the length of the conduction block. This study evaluated the quality of life 15 days after performing this block to treat RTN.

**Methods:** This retrospective study included all patients who, after informed consent, received iterative trigeminal blocks to treat a RTN between 2014 and 2018 in a university hospital. Patients received 0.5% levobupivacaine in combination with clonidine and a corticosteroid (cortivazol or betamethasone according their availability). Data were obtained from patients medical data files and a telephone questionnaire for the SF-12 score. The main criteria of evaluation was the change in quality of life according SF-12 performed at day 15.

**Results:** Twenty-one patients aged 62 ±14 years were included. All patients exhibited RTN after many different clinical treatments according ICHD-3 criteria. Seventy-one per cent of RTN occurred after trauma or surgery. Before receiving blocks, SF-12 physical (SF12-PS) and mental (SF-12 MS) scores reached respectively 35 ± 14 and 29 ± 11. A mean time of 4 ± 5 years elapsed between the occurrence of RTN and nerve blockade. At day 15, SF-12 PS increased by a 3 point mean value and SF-12 MS by 5 points. Approximately half of the patients (55%) were considered as non-responders with a cut-off value of less than 10% variation of their initial SF-12 score. When excluding these patients, SF-12 PS and SF-12 MS were increased by 17 and 9 points respectively. The mean duration of blocks lasted 15 ± 59 days and no severe adverse effects were observed. Patient satisfaction was correlated with increased SF-12 PS (r^2^ = 0.3 p = 0.01) and with the length of analgesia (r^2^ = 0.51 p = 0.001) but not to SF-12 MS variation (p = 0.12).

**Conclusion:** Trigeminal nerve blocks are temporarily effective on pain that may increase the quality of life in responder patients. The reason why some patients are unresponsive to this treatment and why durations in efficacy are so variable remain unsolved. However, in responders, trigeminal nerve blocks seem simple, harmless, not excessively cumbersome and without severe adverse effects.

## Introduction

Chronic pain is a multidimensional illness with a severe negative impact on patient quality of life. Trigeminal neuralgia is a clinical entity involving many etiologies causing a neuropathic pain that is sometimes refractory to all medical treatments^1^. Its incidence reaches 12/100,000 people/year and 1.8/100,000 people /year when neuralgia involves only one branch of the trigeminal nerve^2^. Twenty-five percent of these neuralgias are refractory to carbamazépine^3^, and surgical advice is recommended as first line treatment. Alternative therapies such as peripheral trigeminal blocks may be proposed to patients despite limited scientific evidence^4,5^. According to experts, improvement of functional status and quality of life must be the main criteria to evaluate the utility and effect of loco-regional anesthesia to treat chronic pain^5^. Trigeminal block efficacy on chronic pain is controversial but when effective, its prolonged clinical beneficial effect on pain is far longer than the pharmacological effects of conduction blocks suggesting that local anesthetics may act on the pain mechanism^6^. Indeed this effect may be prolonged for several days to weeks ^7,8,9^. Only one study evaluated the effect of a single trigeminal nerve block (TNB) on quality of life. This study only included 6 patients and did not show a substantial improvement in quality of life 30 and 90 days after performing the block. Moreover, quality of life was a secondary objective of this study^10^. The main objective of the present study was thus to evaluate the effect of TNB on the quality of life 15 days after TNB in patients suffering from trigeminal neuralgia refractory to classically recommended clinical treatment. Secondary objectives were to evaluate the reduction in pain levels at 15 days as well as analgesic consumption, duration of block, and patient satisfaction.

## Methods

This observational, descriptive, monocentric study was retrospectively performed in the postoperative pain clinic of a university hospital in patients receiving TNB between 2014 and 2018. Informed consent was obtained from patients after ethics committee approval (N° ID-RCB 2018-A01979-46) and ClinicalTrials.gov registration (NCT03669744).

Patients were included whenever they suffered from refractory trigeminal neuralgia and were referred from other pain clinics that did not perform such techniques. Patients received one or several blocks between 2014 et 2018 and were excluded when they declined to participate in the study, when they received another type of face block during the previous year, and in the case of insufficient data in the medical file or death at the time of the questionnaire. The telephone questionnaire was completed between July 2018 and March 2019.

During the first consultation, topography of neuralgia, complete neurologic examination and information on the procedure were assessed the by anesthesiologist and pain nurse. After patient consent, blocks were realized in conscious patients at the level of emergence of trigeminal nerve in the supra / infra zygomatic fossa or the supra orbital fossa as described by Pulcini et al.^11^ A combination of 0.5% levobupivacaine, corticosteroid (cortivazol 3.75mg/1ml,or betamethasone 7mg/1ml when available) and 1μg.kg^-1^ clonidine were injected near the nerve trunk after a negative aspiration test. Patients were monitored 30 min following blocks in the post anesthesia room. Pain was evaluated before and 30 minutes after performing the block. Telephone follow-up calls were made 15 days following the last block.

Quality of life was evaluated before and at 15 days following nerve blocks by the SF-12 questionnaire in all patients. A cut-off value of less than 10% variation in their initial SF-12 score defined non-responders. Visual analogic pain score and DN4 scale quantified pain before and after nerve blocks. Patients were instructed to call to schedule a new block when pain reappeared. This time lag defined duration of analgesia. Patient satisfaction, analgesic consumption and adverse effects were evaluated at day 15.

Statistical analyses were done with XLSTAT software. Data obtained between day 0 and day 15 were compared by paired t-test. Linear regression analysis analyzed the link between SF-12 scores and other quantitative data and Cohen Kappa concordance tests. A p-value < 0.05 was considered statistically significant.

## Results

Twenty-five patients received trigeminal nerve blocks between 2014 and 2018. Four patients were excluded: one died before inclusion, one remained unreachable, one refused to participate in the study and the last one had received another face block in the past year (Figure 1). Demographic and clinical data of the 21 remaining patients are given in Table 1 (62 ± 14 years; sex ratio 0.6). Before performing nerve blocks, all patients received an anti-epileptic: 71% received carbamazepine, 76% amitriptyline and 95% an antidepressant. Fifty-two percent took an opioid among which 28% received morphine, fentanyl or oxycodone and 47% tramadol. Two patients had nerve surgery: thermocoagulation in one and micro-vascular decompression in the other. Patients had received an average of 4 TNB at the time of the study.

**Table 1.** General characteristics of patient population: Median (IQR) or Frequency (%).

|  |  |
| --- | --- |
| **Sex ratio (M/F 0.6)** | 21 (100%) |
| **Age (years)** | 62±14 |
| 30 -49 | 4 (19%) |
| 50 - 69 | 14 (67%) |
| ≥ 70 | 3 (14%) |
| **Other Diseases** |  |
| none | 2 (10%) |
| Cardiovascular | 7 (33%) |
| Pulmonary | 2 (10%) |
| Neurological | 4 (20%) |
| Metabolic | 7 (33%) |
| Digestive | 2 (10%) |
| Chronic inflammatory | 3 (14%) |
| Cancers | 2 (10%) |
| Allergy | 4 (20%) |
| Psychiatric and mood | 4 (20%) |
| Other chronic pain | 9 (43%) |
| **Etiology** |  |
| Idiopathic | 1 (5%) |
| Post-surgical | 8 (38%) |
| Post dental care | 7 (33%) |
| Post traumatic | 0 (0%) |
| Post zona | 2 (10%) |
| Neurovascular conflict unoperated | 2 (10%) |
| ENT Cancer | 1 (5%) |
| Intracranial tumor | 1 (5%) |
| **Time to care (years)** | 4±5 |
| ≤ 1 | 2 (10%) |
| 2 – 4 | 9 (43%) |
| 5 – 9 | 6 (28%) |
| ≥ 10 | 4 (20%) |
| **Types of blocks** |  |
| V1 | 4 (20%) |
| V2 | 4 (20%) |
| V3 | 6 (28%) |
| V1+V2 | 4 (20%) |
| V2+V3 | 3 (14%) |

**Figure 1:**
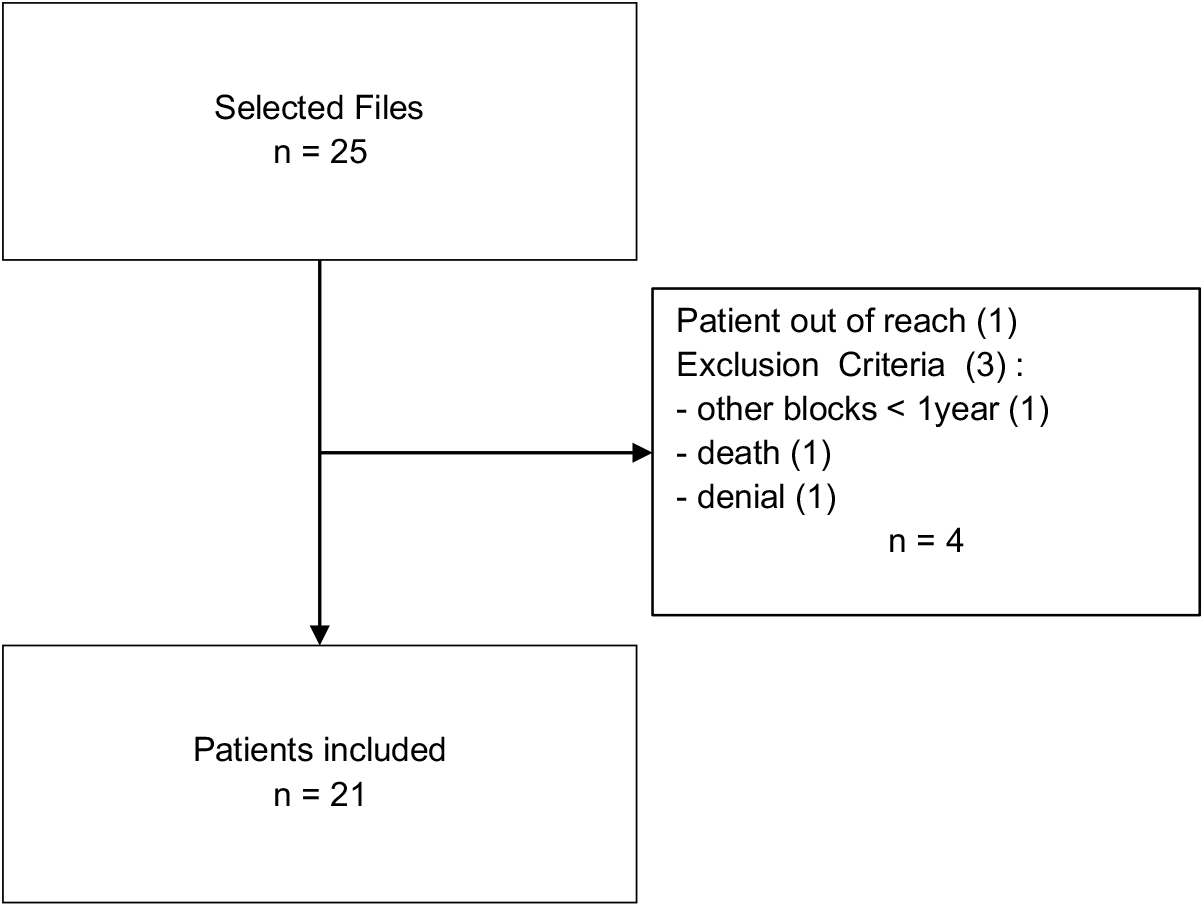
Flow Chart.

SF12 scores are shown in Figure 2. Eleven patients (52%) exhibited less than 10% increase in SF-12 PS and 12 patients (57%) in SF-12 MS. These patients were considered as non-responders. When excluding these non-responders, SF-12 PS increased by 16.6 points [12.1387 - 6.3033], p < 0.01), and SF-12 MS by 9.2 points [27.1476 - 6.0769], p < 0,01). No correlation was observed between SF-12 PS score with sex or age of patients, the time-lag between first symptoms and first block, or anatomical location of pain. SF-12 PS scores variation were correlated with patient satisfaction (r^2^ = 0.30, p = 0.01) but not SF-12 MS (p =0.12).

**Figure 2:**
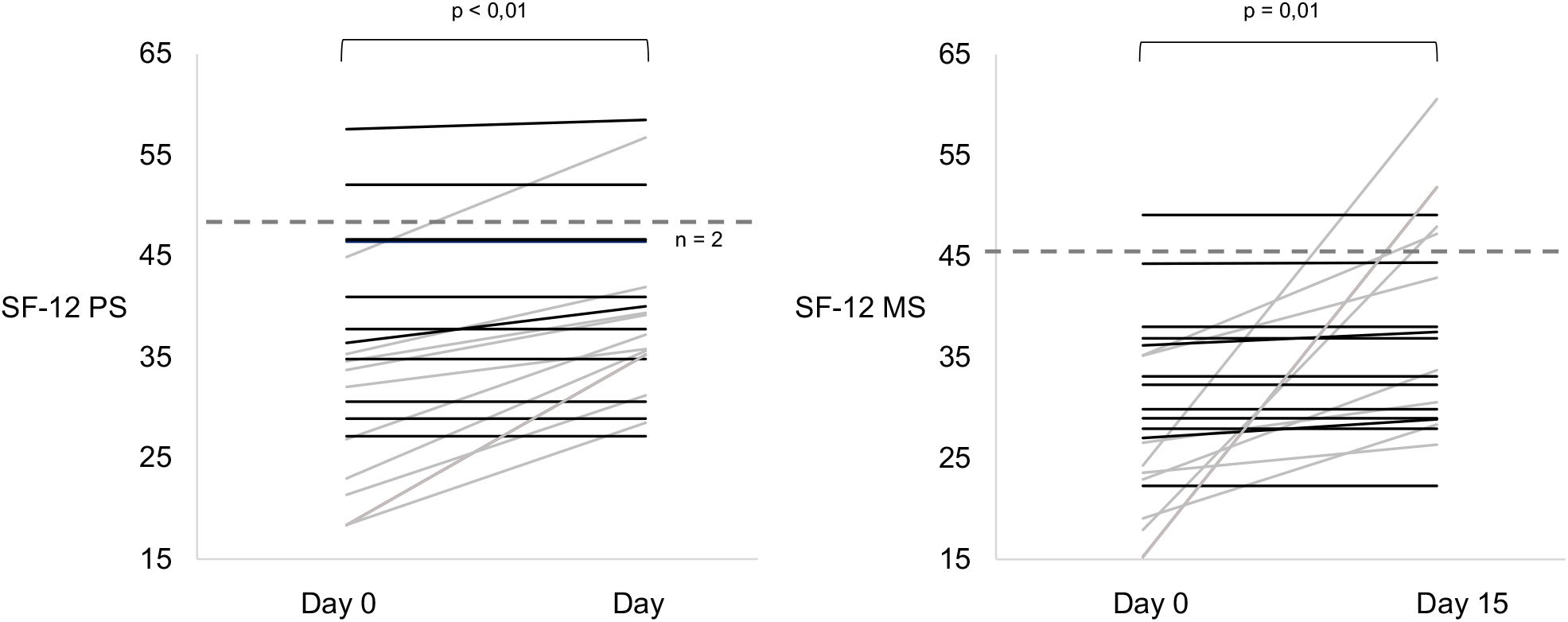
Individual SF-12 physical and mental status scores. Patients with less than 10% improvement are represented in black. Dashed lines represent national mean values of normal patients.

Pain was reduced by a mean of 3 points (t 4.578; IC 95% [1.8145; 4.8522]; p < 0.001). Block efficacy was immediate on the neuropathic pain component with a mean 3 point decrease in the DN4 score (Figure 3). Block mean durations reached 15± 59 days with 1 and 90 days as extreme values. Patient satisfaction was correlated with block duration (r^2^ = 0.51; p < 0.01). Among the 11 responder patients, 9 had a satisfaction score ≥ 7. The 2 others had an unchanged SF-12 MS score. All non-responders had a satisfaction score ≤ 5 (Cohen Kappa concordance 0.81). Analgesic consumption and specifically opioid use was reduced by 61% between the first block and the time of the telephone questionnaire (Figure 4). Six adverse effects were observed: one diplopia, one edema at the puncture site, local dysesthesia on the block territory in one case, 3 cases of pain during puncture. In all cases, these effects were transitory. The block was considered as well tolerated according a visual analog scale of 7/10 rating experience as bad (0/10) to excellent (10/10).

**Figure 3:**
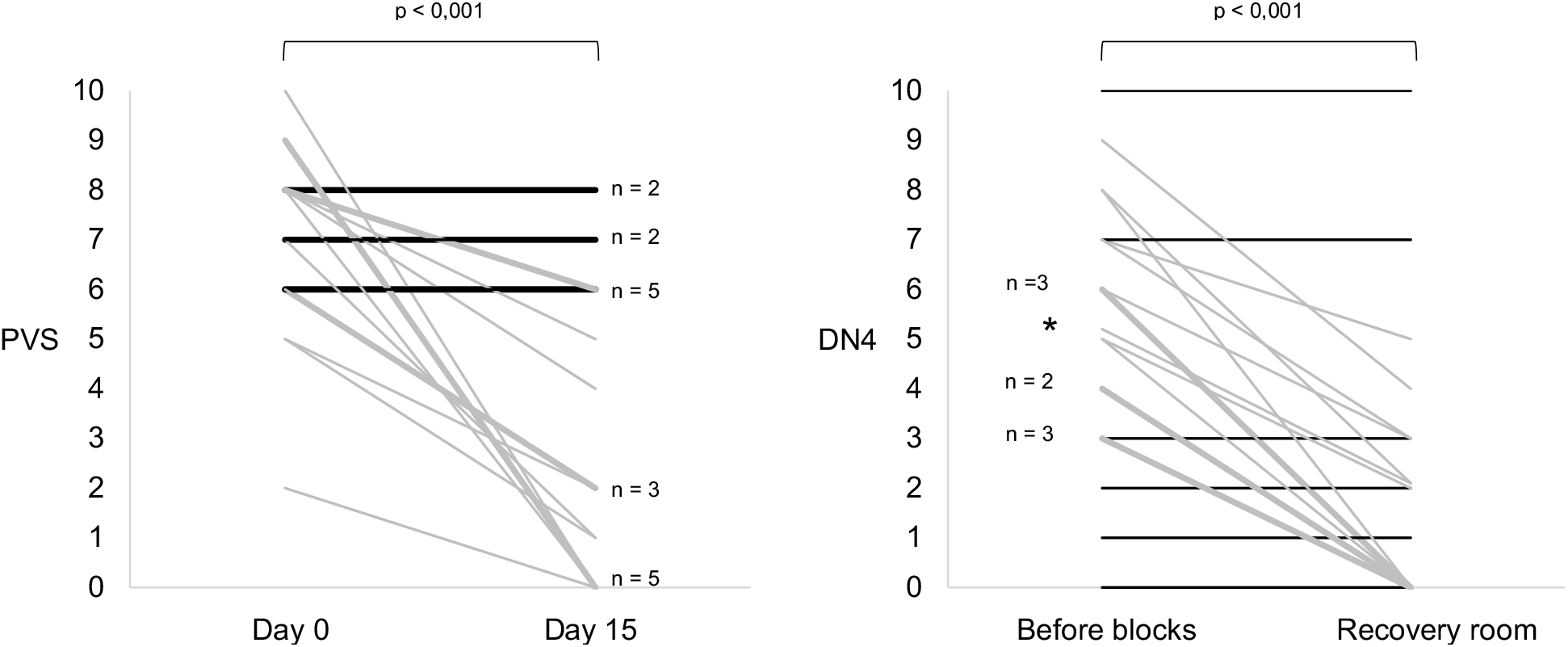
Pain Verbal Scale and DN4 scores. Black lines represent patients with less than 10% improvement.

**Figure 4:**
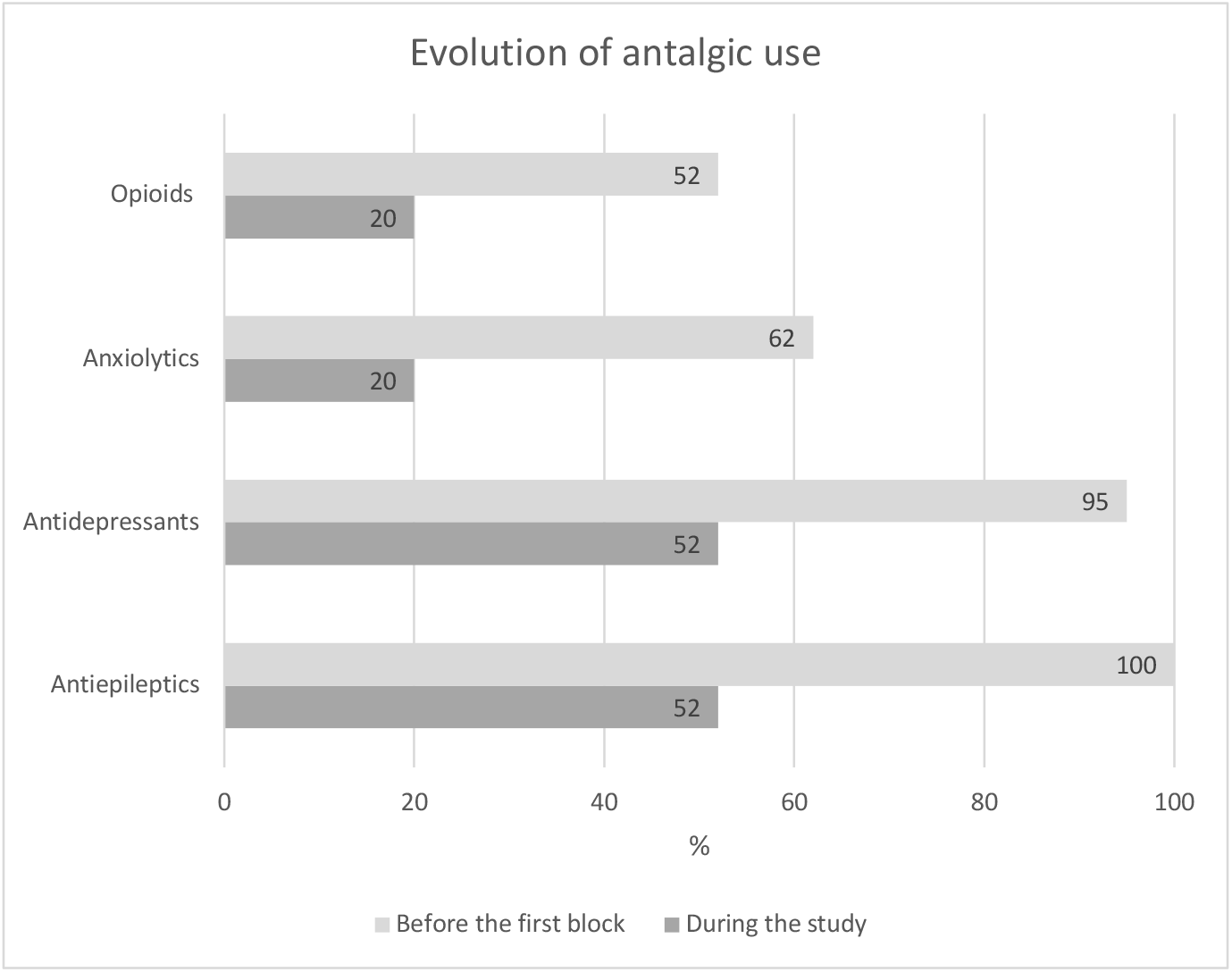
Antalgic use before and after performing single or repeated blocks.

## Discussion

This retrospective monocentric study observed a significant mean improvement in quality of life according the SF-12 scale after TNB in 21 patients suffering from trigeminal neuralgia refractory to usual treatments. However, 50% of patients were non-responders to these blocks. When excluding these patients, the SF-12 PS score improved by 17 points to similar levels of a reference population. Improvement in quality of life was correlated with pain reduction, block efficacy and increased duration and patient satisfaction. Overall background pain therapies were reduced in half of the patients, and opioids in 63 %. Transitory minor adverse effects were observed in six cases but no major effects. These results agreed with the only other study available on quality of life after TNB for refractory trigeminal neuralgia. This previous study was, however, performed in only 6 patients with trigeminal pain of heterogenous origin and used SF-36 quality of life questionnaires. In this latter study, quality of life scores were higher in the group which had already received blocks before receiving the blocks except for the “SF-36 physical role functioning” item. This item improved after blocks similarly to the SF-12 PS of the present study. We chose the SF-12 score because it is easy, rapid and feasible for a telephone interview. The EQ5D score was not performed despite its wide-spread use in the literature because it has a ceiling effect and a potential risk to under-detect small variations in quality of life. Variations in percentage to differentiate responders from non-responders was pragmatic because a minimal improvement threshold after loco-regional anesthesia has not been previously defined. Clinically pertinent SF-12 score improvements are heterogeneous in the literature. For example, in patients with lombalgia, 3 and 4 point variations in SF-12 PS may be considered clinically pertinent with lower values in patients with chronic pain and in those having a better quality of life^12^. In contrast, after knee surgery, 9 and 14 point variations in SF-12 PS and MS are the threshold clinically pertinent values detected and only 1.5 to 2 points the minimal significantly detectable clinical variations in 90% of patients^13^. We considered in this paper that percentage variations were more pertinent than variations expressed as absolute values. Indeed, only a small increase in patients with a poor quality of life had a potentially higher impact than the same absolute variation in patients with a better basal quality of life. In this study, the good concordance between 10% improvement in SF-12 PS and patient satisfaction suggests that this10% threshold was clinically pertinent.

Other factors than pain reduction may explain quality of life improvement or patient satisfaction. Empathic care in patients regarded as incurable or whose pain is disregarded is a major component of patient satisfaction ^14^. In the present study, the lack of contentment or of SF-12 score improvement at day 15 when the TNB is ineffective suggest that this psychological component is not sufficient to explain SF-12 score improvement. This also argues against a placebo effect even if Bischoff et al. observed 40% of responders in the placebo group after ilio/hypogastric nerve blocks^15^. Reducing the number of treatments against pain that may reduce their adverse effects is also a major improvement factor in quality of life. In this study, at the time of the telephone questionnaire, opioid use was reduced by 63% and antiepileptic drugs by 48%.

Querying Pub Med from 2013 with MeSH terms « trigeminal nerve block », « peripheral nerve block », or « trigeminal neuralgia » found mainly isolated clinical cases or series presented in Table 2. Trigeminal neuralgia trigger-zone infiltration has only moderate efficacy on pain levels^16^. In all these papers, patient criteria of selection as well as type of blocks were heterogeneous leading to difficulties to compare with our results. In one study published only as an abstract in 2014, Calot et al.^17^ observed similar results with a 4 point mean VAS after performing face blocks in 44 patients and quality of life improvement in 82% of them without defining the SF-12 performance threshold. In one case Nader et al. in 2015^8^ observed a 50% reduction in pain levels, 15 days after performing a block on the V2 branch in a patient suffering from refractory trigeminal pain for 4 years. More recently, a retrospective study^9^ observed, analgesia lasting between 1 to 8 months after trigeminal nerve blocks in 6 out of 9 patients. In only one randomized study, quality of life was investigated in 13 patients suffering from trigeminal pain^10^. Six patients received TNB using 2% lidocaine injected in the supra/infra orbitary or mandibular foramens. At days 30 and 90, pain attacks were less frequent, pain levels reduced and quality of life improved according to the SF-36 questionnaire.

**Table 2:**
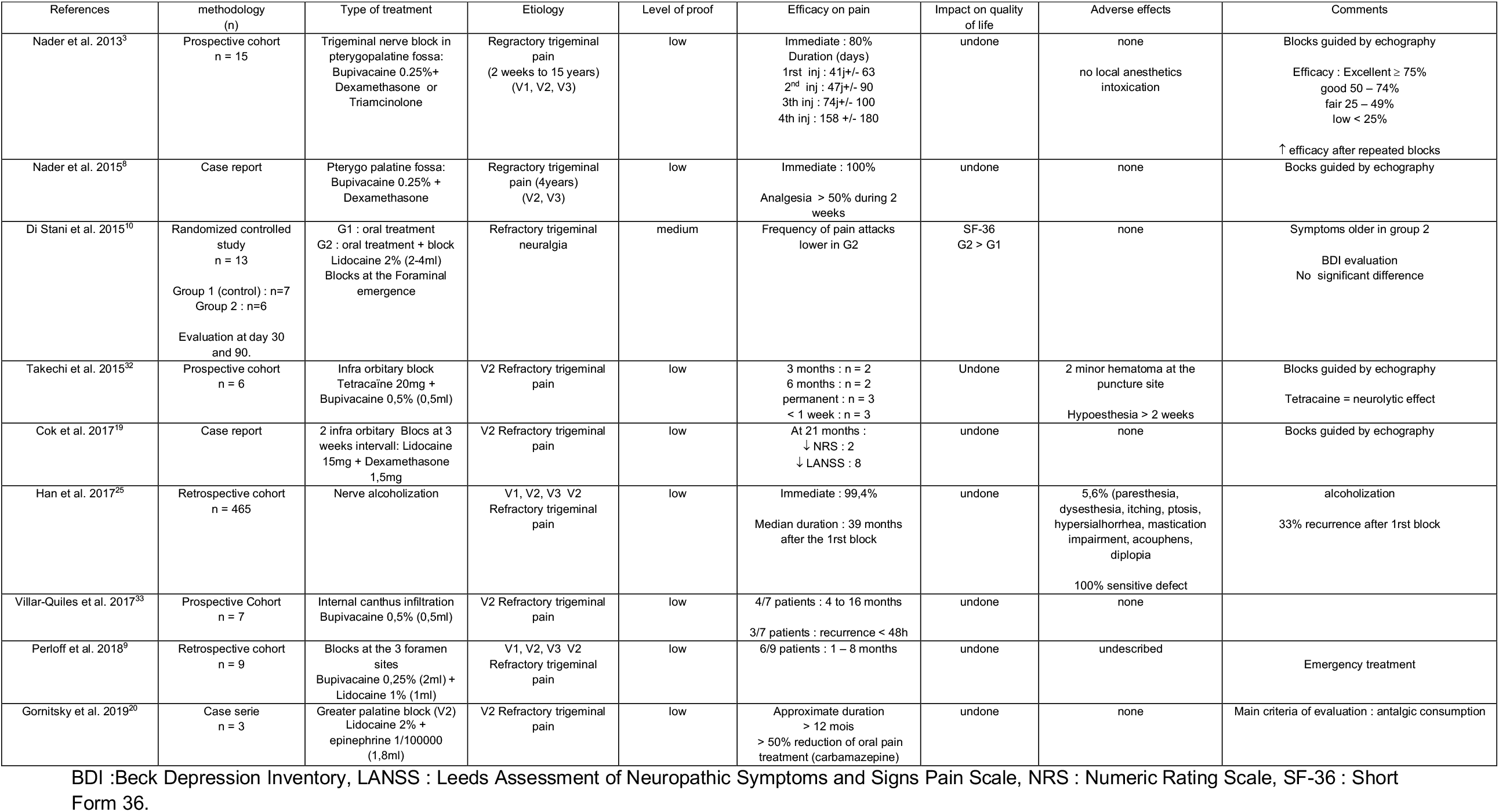
Literature review 2013 – 2019.

| References | methodology<br>(n) | Type of treatment | Etiology | Level of proof | Efficacy on pain | Impact on quality<br>of life | Adverse effects | Comments |
| --- | --- | --- | --- | --- | --- | --- | --- | --- |
| Nader et al. 2013<sup>3</sup> | Prospective cohort<br>n = 15 | Trigeminal nerve block in<br>pterygopalatine fossa:<br>Bupivacaine 0.25%+<br>Dexamethasone or<br>Triamcinolone | Refractory trigeminal<br>pain<br>(2 weeks to 15 years)<br>(V1, V2, V3) | low | Immediate : 80%<br>Duration (days)<br>1st inj : 41j+/- 63<br>2<sup>nd</sup> inj : 47j+/- 90<br>3th inj : 74j+/- 100<br>4th inj : 158 +/- 180 | undone | none<br>no local anesthetics<br>intoxication | Blocks guided by echography<br>Efficacy : Excellent ≥ 75%<br>good 50 – 74%<br>fair 25 – 49%<br>low < 25%<br>↑ efficacy after repeated blocks |
| Nader et al. 2015<sup>8</sup> | Case report | Pterygo palatine fossa:<br>Bupivacaine 0.25% +<br>Dexamethasone | Refractory trigeminal<br>pain (4years)<br>(V2, V3) | low | Immediate : 100%<br>Analgesia > 50% during 2<br>weeks | undone | none | Bocks guided by echography |
| Di Stani et al. 2015<sup>10</sup> | Randomized controlled<br>study<br>n = 13<br>Group 1 (control) : n=7<br>Group 2 : n=6<br>Evaluation at day 30<br>and 90. | G1 : oral treatment<br>G2 : oral treatment + block<br>Lidocaine 2% (2-4ml)<br>Blocks at the Foraminal<br>emergence | Refractory trigeminal<br>neuralgia | medium | Frequency of pain attacks<br>lower in G2 | SF-36<br>G2 > G1 | none | Symptoms older in group 2<br>BDI evaluation<br>No significant difference |
| Takechi et al. 2015<sup>32</sup> | Prospective cohort<br>n = 6 | Infra orbitary block<br>Tetracaine 20mg +<br>Bupivacaine 0,5% (0,5ml) | V2 Refractory trigeminal<br>pain | low | 3 months : n = 2<br>6 months : n = 2<br>permanent : n = 3<br>< 1 week : n = 3 | Undone | 2 minor hematoma at the<br>puncture site<br>Hypoesthesia > 2 weeks | Blocks guided by echography<br>Tetracaine = neurolytic effect |
| Cok et al. 2017<sup>19</sup> | Case report | 2 infra orbitary Blocs at 3<br>weeks intervall: Lidocaine<br>15mg + Dexamethasone<br>1,5mg | V2 Refractory trigeminal<br>pain | low | At 21 months :<br>↓ NRS : 2<br>↓ LANSS : 8 | undone | none | Bocks guided by echography |
| Han et al. 2017<sup>25</sup> | Retrospective cohort<br>n = 465 | Nerve alcoholization | V1, V2, V3 V2<br>Refractory trigeminal<br>pain | low | Immediate : 99,4%<br>Median duration : 39 months<br>after the 1st block | undone | 5,6% (paresthesia,<br>dysesthesia, itching, ptosis,<br>hypersialhorrea, mastication<br>impairment, acouphens,<br>diplopia<br>100% sensitive defect | alcoholization<br>33% recurrence after 1st block |
| Villar-Quiles et al. 2017<sup>33</sup> | Prospective Cohort<br>n = 7 | Internal canthus infiltration<br>Bupivacaine 0,5% (0,5ml) | V2 Refractory trigeminal<br>pain | low | 4/7 patients : 4 to 16 months<br>3/7 patients : recurrence < 48h | undone | none |  |
| Perloff et al. 2018<sup>9</sup> | Retrospective cohort<br>n = 9 | Blocks at the 3 foramen<br>sites<br>Bupivacaine 0,25% (2ml) +<br>Lidocaine 1% (1ml) | V1, V2, V3 V2<br>Refractory trigeminal<br>pain | low | 6/9 patients : 1 – 8 months | undone | undescribed | Emergency treatment |
| Gornitsky et al. 2019<sup>20</sup> | Case serie<br>n = 3 | Greater palatine block (V2)<br>Lidocaine 2% +<br>epinephrine 1/100000<br>(1,8ml) | V2 Refractory trigeminal<br>pain | low | Approximate duration<br>> 12 mois<br>> 50% reduction of oral pain<br>treatment (carbamazepine) | undone | none | Main criteria of evaluation : antalgic consumption |

BDI :Beck Depression Inventory, LANSS : Leeds Assessment of Neuropathic Symptoms and Signs Pain Scale, NRS : Numeric Rating Scale, SF-36 : Short Form 36.

The combination used to perform the blocks in this study included 3 drugs: a long-lasting local anesthetic (0.5% levobupivacaine), a long-lasting corticosteroid and clonidine. The choice and doses of the drugs were based on routine practice, in order to prolong the duration of locoregional anesthesia and potentially act on chronic pain mechanisms as previously published^18^. The most frequently used treatment combines a local anesthetic and a long lasting corticosteroid (usually dexamethasone)^3,8,19^. In many cases, when using very short action lidocaine, blocks lasted far longer than in the present study in which a long-acting local anesthetic was used. For example Cok et al.^19^ obtained a 21 month analgesia after a single infra-orbitary nerve block with 15 mg lidocaine and 1.5mg dexamethasone. In 2019, Gornitsky et al.^20^ similarly observed several months pain relief in 3 patients after spheno-palatin nerve blockade with adrenaline-lidocaine. In the present study, block durations were not as long and were more variable when using a long lasting local anesthetic. Lidocaine neurotoxicity might explain these differences.

Corticosteroids are frequently used for chronic pain treatment either by local or perineural infiltration. They are effective for neuropathic pain relief ^21^ and for prolonging analgesia after peripheral nerve blocks ^22,23^. Complete proof of their safety and absence of neurotoxicity are lacking. A recent anatomopathological study in mice, did not find Wallerian degeneracy nor perineural inflammation when dexamethasone was used alone or in combination with ropivacaine^24^. Finally, in 2 meta analyses including 155 et 801 patients, no adverse effects were observed after perineural corticosteroid injection^25,26^.

Clonidine perineural use is less common for chronic pain management. For example, Maranto et al.^18^, consider that potential benefits are higher than risks. Clonidine use is widely used for acute pain management with loco-regional anesthesia to increase and deepen analgesia^27^ and prevent neuropathic pain^28^.

Regardless of the combination used, long or short acting local anesthetics with or without adjuvants, analgesia is far longer than the sole conduction block induced by local anesthetics. Breaking the vicious circle of chronic pain with locoregional anesthesia may partly explain the success of nerve blocks in chronic pain management.

Other therapeutic options including microvascular decompression and thermocoagulation are equally effective^29^ but of longer duration ^30^. However, they require general anesthesia and sometimes repeated procedures to obtain optimal efficacy. Thirty to 45% of patients must continue medical treatment after surgery^31^. Prolonged adverse effects can be observed such as 15 to 50% sensory impairment ^31^ and, although rare, mortality after microvascular decompression ^30^. After trigeminal alcoholization, 39 months relief was observed in a 465 patient retrospective cohort ^29^ but the complications rate reached 8.6% and all patients had some sort of sensory impairment. Trigeminal nerve blocks can be performed in an ambulatory setting, there by avoiding general anesthesia and its complications and are well tolerated as measured in this study.

This study has several bias and limits. Trigeminal neuralgia is rare and 25% of patients can be refractory to a well conducted clinical treatment^23^. This may explain why locoregional anesthesia efficacy is not fully demonstrated with regards to the low number of patients in the previous studies. Similarly, we included a limited number of patients leading to limited statistical power. Selection bias is unavoidable because 71% of patients were referred after ENT, dentistry oy neurosurgery. In the present study population, post-traumatic trigeminal pain was over represented. The retrospective nature of the study engenders information and memorisation bias. Twenty per cent of patients in the present study had a depressive syndrome. The lack of correlation between the SF-12 PS and SF-12 MS scores before or after TNB confirms that performing the block is not sufficient and that specific psychological treatment is required. Specific depression-anxiety scales were not obtained for all patients and we did not correlate these scales with SF-12 MS scores.

## Conclusion

This study demonstrated improved quality of life in patients suffering from refractory trigeminal pain fifteen days after specific nerve blocks. Half of patients were responders to this anesthetic technique and their quality of life was restored to similar levels compared to a reference population. This technique is simple, harmless and well accepted by patients. Further prospective multicentric studies are needed to confirm these results on quality of life, determine why some patients are unresponsive to this technique and to identify the best combination of drugs to inject in proximity to the nerves.

## Data Availability

No link, only in Excel files.

## Acknowledgements

The authors thank Arthur Baïsse for his critical review, Chantal and Laurent for their availability, Dr Jeanne Cook-Moreau for translation assistance.

## References

1. Maarbjerg S, Di Stefano G, Bendtsen L, Cruccu G. Trigeminal neuralgia – diagnosis and treatment. Cephalalgia. juin 2017;37(7):648–57.

2. Koopman JSHA, Dieleman JP, Huygen FJ, de Mos M, Martin CGM, Sturkenboom MCJM. Incidence of facial pain in the general population: Pain. déc 2009;147(1):122–7.

3. Nader A, Kendall MC, De Oliveria GS, Chen JQ, Vanderby B, Rosenow JM, et al. Ultrasound-guided trigeminal nerve block via the pterygopalatine fossa: an effective treatment for trigeminal neuralgia and atypical facial pain. Pain Physician. oct 2013;16(5):E537–545.

4. Cruccu G, Truini A. Refractory Trigeminal Neuralgia: Non-Surgical Treatment Options. CNS Drugs. févr 2013;27(2):91–6.

5. Beloeil H, Viel é., Navez M-L, Fletcher D, Peronnet D. Techniques analgésiques locorégionales et douleur chronique. Annales Françaises d’Anesthésie et de Réanimation. avr 2013;32(4):275–84.

6. Vlassakov KV, Narang S, Kissin I. Local Anesthetic Blockade of Peripheral Nerves for Treatment of Neuralgias: Systematic Analysis. Anesthesia & Analgesia. juin 2011;112(6):1487–93.

7. Arnér S, Lindblom U, Meyerson BA, Molander C. Prolonged relief of neuralgia after regional anesthetic blocks. A call for further experimental and systematic clinical studies: Pain. déc 1990;43(3):287–97.

8. Nader A, Bendok BR, Prine JJ, Kendall MC. Ultrasound-Guided Pulsed Radiofrequency Application via the Pterygopalatine Fossa: A Practical Approach to Treat Refractory Trigeminal Neuralgia. Pain Physician. juin 2015;18(3):E411–415.

9. Perloff MD, Chung JS. Urgent care peripheral nerve blocks for refractory trigeminal neuralgia. The American Journal of Emergency Medicine. nov 2018;36(11):2058–60.

10. Stani FD, Ojango C, Dugoni D, Lorenzo LD, Masala S, Delfini R, et al. Combination of pharmacotherapy and lidocaine analgesic block of the peripheral trigeminal branches for trigeminal neuralgia: a pilot study. Arq Neuro-Psiquiatr. août 2015;73(8):660–4.

11. Pulcini A, Guerin J-P, Sibon S, Balaguer T, Ichai C. Blocs de la face. EMC - Anesthésie-Réanimation. janv 2007;4(3):1–14.

12. Díaz-Arribas MJ, Fernández-Serrano M, Royuela A, Kovacs FM, Gallego-Izquierdo T, Ramos-Sánchez M, et al. Minimal Clinically Important Difference in Quality of Life for Patients With Low Back Pain: SPINE. déc 2017;42(24):1908–16.

13. Clement ND, Weir D, Holland J, Gerrand C, Deehan DJ. Meaningful changes in the Short Form 12 physical and mental summary scores after total knee arthroplasty. The Knee. août 2019;26(4):861–8.

14. Sturgeon J. Psychological therapies for the management of chronic pain. PRBM. avr 2014;115.

15. Bischoff JM, Koscielniak-Nielsen ZJ, Kehlet H, Werner MU. Ultrasound-Guided Ilioinguinal/Iliohypogastric Nerve Blocks for Persistent Inguinal Postherniorrhaphy Pain: A Randomized, Double-Blind, Placebo-Controlled, Crossover Trial. Anesthesia & Analgesia. juin 2012;114(6):1323–9.

16. Lemos L, Flores S, Oliveira P, Almeida A. Gabapentin Supplemented With Ropivacain Block of Trigger Points Improves Pain Control and Quality of Life in Trigeminal Neuralgia Patients When Compared With Gabapentin Alone: The Clinical Journal of Pain. janv 2008;24(1):64–75.

17. Calot C, Laforge E, Lasserre A, Nouette-Gaulain K. Efficacité de l’anesthésie locorégionale (ALR) dans les algies faciales chroniques. Annales Françaises d’Anesthésie et de Réanimation. sept 2014;33:A288–9.

18. Maranto CJ, Strickland NR, Goree JH. Combined Superficial and Deep Serratus Plane Block With Bupivacaine, Dexamethasone, and Clonidine in the Treatment of a Patient With Postmastectomy Pain Syndrome: A Case Report. A & A Practice. nov 2018;11(9):236–7.

19. Cok OY, Deniz S, Eker HE, Oguzkurt L, Aribogan A. Management of isolated infraorbital neuralgia by ultrasound-guided infraorbital nerve block with combination of steroid and local anesthetic. Journal of Clinical Anesthesia. févr 2017;37:146–8.

20. Gornitsky M, Elsaraj SM, Canie O, Mohit S, Velly AM, Schipper HM. Greater palatine block for V2 trigeminal neuralgia: Case report. Special Care in Dentistry. mars 2019;39(2):208–13.

21. Eker HE, Cok OY, Aribogan A, Arslan G. Management of Neuropathic Pain with Methylprednisolone at the Site of Nerve Injury. Pain Medicine. mars 2012;13(3):443–51.

22. Heesen M, Klimek M, Imberger G, Hoeks SE, Rossaint R, Straube S. Co-administration of dexamethasone with peripheral nerve block: intravenous vs perineural application: systematic review, meta-analysis, meta-regression and trial-sequential analysis. British Journal of Anaesthesia. févr 2018;120(2):212–27.

23. Chong MA, Berbenetz NM, Lin C, Singh S. Perineural Versus Intravenous Dexamethasone as an Adjuvant for Peripheral Nerve Blocks: A Systematic Review and Meta-Analysis. Regional Anesthesia and Pain Medicine. 2017;42(3):319–26.

24. Marty P, Bennis M, Legaillard B, Cavaignac E, Ferre F, Lebon J, et al. A New Step Toward Evidence of In Vivo Perineural Dexamethasone Safety: An Animal Study. Regional Anesthesia and Pain Medicine. avr 2017;1.

25. Khan JS, Rai A, Sundara Rajan R, Jackson TD, Bhatia A. A scoping review of perineural steroids for the treatment of chronic postoperative inguinal pain. Hernia. juin 2016;20(3):367–76.

26. Choi S, Rodseth R, McCartney CJL. Effects of dexamethasone as a local anaesthetic adjuvant for brachial plexus block: a systematic review and meta-analysis of randomized trials. British Journal of Anaesthesia. mars 2014;112(3):427–39.

27. Naja Z, Al-Tannir M, El-Rajab M, Ziade F, Daher Y, Khatib H, et al. The Effectiveness of Clonidine-Bupivacaine Repeated Nerve Stimulator-guided Injection in Piriformis Syndrome: The Clinical Journal of Pain. mars 2009;25(3):199–205.

28. Mohamed SA-B, Abdel-Ghaffar HS. Effect of the addition of clonidine to locally administered bupivacaine on acute and chronic postmastectomy pain. Journal of Clinical Anesthesia. févr 2013;25(1):20–7.

29. Han KR, Chae YJ, Lee JD, Kim C. Trigeminal nerve block with alcohol for medically intractable classic trigeminal neuralgia: long-term clinical effectiveness on pain. International Journal of Medical Sciences. 2017;14(1):29–36.

30. Almeida A, Lemos, Alegria, Oliveira, Oliveira J, Machado. Pharmacological versus microvascular decompression approaches for the treatment of trigeminal neuralgia: clinical outcomes and direct costs. JPR. août 2011;233.

31. Hitchon PW, Holland M, Noeller J, Smith MC, Moritani T, Jerath N, et al. Options in treating trigeminal neuralgia: Experience with 195 patients. Clinical Neurology and Neurosurgery. oct 2016;149:166–70.

32. Takechi K, Konishi A, Kikuchi K, Fujioka S, Fujii T, Yorozuya T, et al. Real-time ultrasound-guided infraorbital nerve block to treat trigeminal neuralgia using a high concentration of tetracaine dissolved in bupivacaine. Scandinavian Journal of Pain. 1 janv 2015;6(1):51–4.

33. Villar-Quiles R-N, García-Moreno H, Mayo D, Gutiérrez-Viedma Á, Ramos M-I, Casas-Limón J, et al. Infratrochlear neuralgia: A prospective series of seven patients treated with infratrochlear nerve blocks. Cephalalgia. mars 2018;38(3):585–91.

